# AMELIE 3: Fully Automated Mendelian Patient Reanalysis at Under 1 Alert per Patient per Year

**DOI:** 10.1101/2020.12.29.20248974

**Authors:** Johannes Birgmeier, Ethan Steinberg, Ethan E. Bodle, Cole A. Deisseroth, Karthik A. Jagadeesh, Jennefer N. Kohler, Devon Bonner, Shruti Marwaha, Julian A. Martinez-Agosto, Stan Nelson, Christina G. Palmer, Joy D. Cogan, Rizwan Hamid, Joan M. Stoler, Joel B. Krier, Jill A. Rosenfeld, Paolo Moretti, David R. Adams, Vandana Shashi, Elizabeth A. Worthey, Christine M. Eng, Euan A. Ashley, Matthew T. Wheeler, Undiagnosed Diseases Network, Peter D. Stenson, David N. Cooper, Jonathan A. Bernstein, Gill Bejerano

## Abstract

**Background:** Many thousands of patients with a suspected Mendelian disease have their exomes/genomes sequenced every year, but only about 30% receive a definitive diagnosis. Since a novel Mendelian gene-disease association is published on average every business day, thousands of undiagnosed patient cases could receive a diagnosis each year if their genomes were regularly compared to the latest literature. With millions of genomes expected to be sequenced for rare disease analysis by 2025, and considering the current publication rate of 1.1 million new articles per annum in PubMed, manually reanalyzing the growing cases of undiagnosed patients is not sustainable.

**Methods:** We describe a fully automated reanalysis framework for patients with suspected, but undiagnosed, Mendelian disorders. The presented framework was tested by automatically parsing all ∼100,000 newly published peer reviewed papers every month and matching them on genotype and phenotype with all stored undiagnosed patients. If a new article contains a possible diagnosis for an undiagnosed patient, the system provides notification. We test the accuracy of the automatic reanalysis system on 110 patients, including 61 with available trio data.

**Results:** Even when trained only on older data, our system identifies 80% of reanalysis diagnoses, while sending only 0.5-1 alerts per patient per year, a 100-1,000-fold efficiency gain over manual literature surveillance of equivalent yield.

**Conclusion:** We show that automatic reanalysis of patients with suspected Mendelian disease is feasible and has the potential to greatly streamline diagnosis. Our system is not intended to replace clinical judgment. Rather, clinical diagnostic services could greatly benefit from a modest re-allocation of time from manual literature exploration to review of automated reanalysis alerts. Our system additionally supports a new paradigm for medical IT systems: proactive, continuously learning and consequently able to autonomously identify valuable insights as they emerge in digital health records. We have launched automated patient reanalysis, trained on the latest data, with user accounts and daily literature updates at https://AMELIE.stanford.edu.

## Introduction

Severe genetic diseases affect tens of thousands of infants born every year worldwide. Many Mendelian conditions such as intellectual disability are diagnosed later in life for a total estimate of 0.5-1% of the 7.8 billion world population^1,2^. Millions of such patients are projected to be sequenced over the next few years^3^. Currently, for an estimated 30% of patients^4^ with a presumed Mendelian disease, a definitive diagnosis is arrived at immediately after exome sequencing^5^. Conversely, 70% of patients do not receive a diagnosis (for a variety of reasons^6^). However, approximately 250 novel gene-disease associations are identified every year^6–8^. Reanalysis of exomes of patients with previously undiagnosable genetic conditions results in a significant fraction (4%-30%) of these cases becoming diagnosable in a period of 1 to 5 years after the initial negative analysis^6,9–17^. PubMed grows by over 1 million publications each year. Thus, the lack of capacity^18^ to regularly reassess non-diagnostic clinical exome or genome sequencing in the light of newly published literature necessarily results in delayed diagnoses.

We have previously developed AMELIE^19–21^ (<u>A</u>utomatic <u>ME</u>ndelian <u>LI</u>terature <u>E</u>valuation), a natural language processing and machine learning framework that automatically analyzes literature about Mendelian diseases and matches it to patients with undiagnosed Mendelian diseases to prioritize candidate causative genes in the patients’ genomes. Here, we adapted the use of AMELIE to perform continuous reanalysis of undiagnosed patients with suspected Mendelian disease. The AMELIE-based reanalysis framework automatically compares *all* new literature to *all* undiagnosed patients and notifies clinicians (or diagnosticians; we use these interchangeably here) about newly published, likely diagnostic articles. To estimate the diagnostic rate and clinician burden of the reanalysis system, we performed a “time machine” experiment: first, we trained the reanalysis system only on Mendelian disease data available until December 2011. Subsequently, we assembled a cohort of 110 Mendelian singleton patients, of which 61 also had trio sequencing data available, who gradually became diagnosable after January 2012. Using this system, we performed an automatic reanalysis experiment in monthly intervals from 2012 to 2018, demonstrating a high diagnostic yield at very low clinician burden.

## Methods

### AMELIE-based automatic reanalysis

The automatic reanalysis framework presented here takes as input exome or genome sequencing data and a (manually or automated ClinPhen^22^-created) list of phenotypic abnormalities per patient. User-parameterized filtering of exome or genome sequencing data reveals a list of patient variants that are rare (e.g., ≤0.5% minor allele frequency^23^) in the general population and hence potentially disease-causing. These are termed “candidate causative” variants. After sequencing, the patient’s candidate causative variants are analyzed for the presence of causative mutations using all knowledge available at the time. If the patient cannot be diagnosed shortly after sequencing, the patient’s relevant data (minimally consisting of a list of candidate causative variants and a list of phenotypic abnormalities observed in the patient) are added to a database of undiagnosed patients. Each patient is then reanalyzed automatically at monthly intervals until a diagnosis is successfully identified (Figure 1).

**Figure 1.**
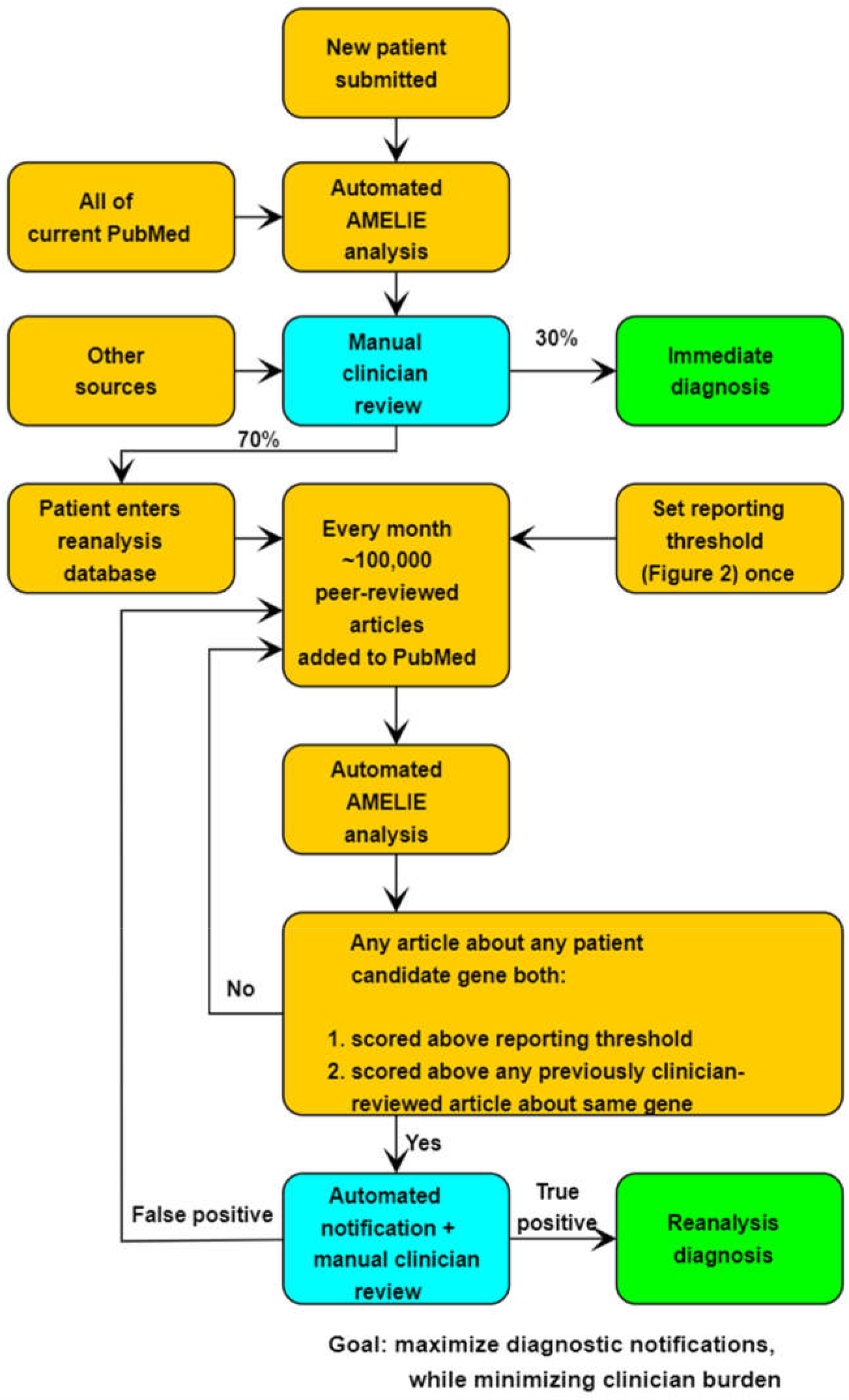
Automatic reanalysis of patients with undiagnosed Mendelian diseases. After sequencing, clinicians examine the automated AMELIE analysis in search of a diagnosis. If a diagnosis is not available (currently in ∼70% of all cases), the patient’s information is entered into a reanalysis database. Every month, AMELIE matches all newly published literature against every patient candidate causative variant and phenotypes to seek new diagnoses. If a newly published article is flagged as being possibly diagnostic, it is reviewed by clinicians, resulting in either diagnosis or continuation of AMELIE-based automatic reanalysis. See example, reanalysis notifications in Table 3.

#### AMELIE

AMELIE^21^ performs two tasks: (1) automatically discovers and parses literature about Mendelian diseases to construct an “AMELIE knowledgebase”, and (2) estimates the likelihood that a given article contains a diagnosis for a patient through an “AMELIE classifier”. Here we build a computational framework around AMELIE that performs automatic reanalysis of undiagnosed patients with suspected Mendelian disease (Figure 1). For a detailed description of AMELIE, see Supplementary Methods and ref. ^21^.

#### AMELIE knowledgebase

The AMELIE knowledgebase is automatically constructed from articles about Mendelian diseases. Briefly, AMELIE knowledgebase construction is performed using a series of machine-learning classifiers^21^ operating on text data. First, all PubMed abstracts available (30+ million currently) are classified in terms of their likelihood to discuss monogenic diseases. The full-text articles of potentially relevant abstracts are retrieved directly from the publishers. From each article’s full text, disease-causing genes and resulting clinical phenotypes are extracted. Mentioned genetic variants are retrieved using AVADA^24^. In addition, a set of full-text classifiers assign scores to each article indicating whether it is most likely to be about a dominant or a recessive disease, and about protein-truncating (frameshift indel, stopgain, splicing) pathogenic variants or non-truncating (missense, nonframeshift indel) pathogenic variants. Information about mentioned phenotypic abnormalities, disease-causing genes, and disease inheritance modes, are extracted from these full text articles into the knowledgebase.

#### AMELIE classifier

The AMELIE classifier estimates the likelihood that a given article contains a diagnosis for a particular patient. Given an article *A*, a patient’s list of phenotypic abnormalities *P*, and a gene *G* containing candidate causative variants in the patient’s genome, the AMELIE classifier^21^ returns a diagnostic probability score between 0 and 100 (low to high) indicating how well the article *A* explains the patient’s phenotypes *P* in light of the patient-specific variants in gene *G*.

#### Automatic reanalysis using AMELIE

The automatic reanalysis framework takes a single parameter as input, termed “notification threshold”, a number (score) between 0 and 100. When a new article *A* about a disease-causing gene *G* is published and added to the AMELIE knowledgebase, the AMELIE classifier compares all known undiagnosed patients with a candidate causative variant in *G* to the article *A* and automatically sends a notification about the article if our “notification criterion” applies. We define the “notification criterion” as (1) article *A*’s diagnostic probability score is greater than or equal to the (global) notification threshold, and (2) article *A*’s diagnostic probability score is greater than or equal to the diagnostic probability score of previously published articles about the candidate gene *G* for the undiagnosed patient.

Patients who are successfully diagnosed after such notifications are removed from the database of undiagnosed patients. If a notification sent by the automatic reanalysis framework contains an article that, after clinician review, enables patient diagnosis, the notification is counted as “diagnostic”, or a “true positive”; if not, it is considered a “false positive” (Figure 1).

### Patients

To retrospectively test AMELIE-based automatic reanalysis, we assembled a cohort of 110 diagnosed patients with diseases where the causative gene was first published between January 2012 and May 2018 (Table 1, Supplementary Table S1). Patient data was obtained from the Deciphering Developmental Disorders (DDD) project^25^, the clinical genetics service at Stanford Children’s Health (SCH), and the Undiagnosed Diseases Network (UDN)^26^. From these sources, we included all available patients with a single causative gene disease diagnosis for which the first supporting literature appeared after January 2012; had available exome or genome sequencing data containing the causative variant(s); and a list of clinician-noted or ClinPhen^22^-extracted phenotypes (Supplementary Methods). De-identified data from the DDD project were accessed via the European Genome-Phenome Archive^27^ (study EGAS00001000775). As applicable to the participating patients, the study protocol was reviewed and approved by the Stanford University Institutional Review Board (IRB) and the central IRB at the NIH National Human Genome Research Institute for the Undiagnosed Diseases Network. Written informed consent was obtained from all participants. For each of the 110 patients, a clinician reviewed the literature about the patient’s disease and manually identified a subset of articles, each with sufficient information to diagnose the case. The year and month in which the first article linking the patient’s disease to the patient’s causative gene was published were tagged as the patient’s earliest possible date of literature-based diagnosis.

**Table 1.** Clinical characteristics of patient cohort.

|  | Singletons | Trios subset |
| --- | --- | --- |
| **Number of patients** | 110 | 61 |
| **Male sex - number (percent)** | 47 (42.7%) | 24 (39.3%) |
| **Phenotypic abnormalities in ... - number (percent)** |  |  |
| Blood and blood-forming tissues | 6 (5.5%) | 5 (8.2%) |
| Breast | 6 (5.5%) | 4 (6.6%) |
| Cardiovascular system | 25 (22.7%) | 16 (26.2%) |
| Connective tissue | 17 (15.5%) | 11 (18.0%) |
| Digestive system | 35 (31.8%) | 21 (34.4%) |
| Ear | 23 (20.9%) | 17 (27.9%) |
| Endocrine system | 15 (13.6%) | 12 (19.7%) |
| Eye | 38 (34.5%) | 27 (44.3%) |
| Genitourinary system | 17 (15.5%) | 6 (9.8%) |
| Growth | 34 (30.9%) | 18 (29.5%) |
| Head or neck | 66 (60.0%) | 40 (65.6%) |
| Immune system | 10 (9.1%) | 7 (11.5%) |
| Integument | 36 (32.7%) | 21 (34.4%) |
| Limbs | 42 (38.2%) | 27 (44.3%) |
| Metabolism/homeostasis | 20 (18.2%) | 12 (19.7%) |
| Musculature | 47 (42.7%) | 31 (50.8%) |
| Neoplasm | 6 (5.5%) | 3 (4.9%) |
| Nervous system (incl. intellectual disability) | 102 (92.7%) | 58 (95.1%) |
| Prenatal development or birth | 22 (20.0%) | 14 (23.0%) |
| Respiratory system | 23 (20.9%) | 14 (23.0%) |
| Skeletal system | 66 (60.0%) | 39 (63.9%) |
| Voice | 2 (1.8%) | 2 (3.3%) |
| **Number of distinct HPO\* phenotypic abnormalities across all patients** | 715 | 558 |
| **Median number of HPO\* phenotypic abnormalities per patient** | 6 | 8 |
| **Number of distinct OMIM\*\* diseases (1 per patient) across all patients** | 74 | 44 |
| **Genetic characteristics** |  |  |
| Median number of candidate causative variants*** per patient | 312 | 25 |
| Median number of candidate causative genes**** per patient | 274 | 20 |
| Number of distinct causative genes across all patients | 74 | 44 |
| *Human Phenotype Ontology (HPO), a database organizing human phenotypic abnormalities |  |  |
| **Online Mendelian Inheritance in Man (OMIM), a database organizing Mendelian diseases |  |  |
| ***Patient variants occurring at $\leq 0.5\%$ allele frequency in a large healthy control cohort (see Supplementary Methods) | | |
| ****Genes containing at least one candidate causative variant |  |  |

We defined candidate causative variants in singleton patient genomes as rare (≤0.5% minor allele frequency in a large healthy control cohort^23^), non-silent exonic or core splice-site variants in protein-coding genes. For 61 of the 110 test patients, exome or genome sequencing data of 2 of the patient’s unaffected relatives (usually parents) were available and the patient’s causative variants were not identically observed in an unaffected relative. For trio patients, candidate variants were further filtered by segregation with the disease in the family (Table 1, Supplementary Table S1).

### Experimental design

For our time machine experiment, we built a version of the AMELIE knowledgebase and trained all machine learning components using only article data from 2011 or before. We then ran this AMELIE classifier, in monthly steps, on all PubMed data from January 2012 through May 2018, noting every notification generated at different notification thresholds (Figure 1).

### Performance Measures

We define the *number of diagnosed patients* as the number of test cohort patients who received a diagnostic notification within the experiment timeframe. The *wait time for diagnosis after publication of the first diagnostic article* is the number of months between the publication of the first diagnostic article and the sending of a diagnostic notification by AMELIE.

In a typical undiagnosed patient set, only a small fraction of patients become diagnosable every year^6,9–17^. Since our test patient cohort consists only of patients who become diagnosable within the experiment timeframe, reporting the number of false positives per diagnostic notification purely from the test cohort data would underestimate the number of false positive notifications per diagnostic notification in a cohort including patients not diagnosable before May 2018. We conducted a meta-analysis of manual reanalysis studies of undiagnosed patients with suspected Mendelian disease^6,9–15^. For each study, we collected the total number of patients, the number of patients receiving a reanalysis diagnosis due to updated literature (rather than other factors like improved variant calling pipelines), and the reanalysis timeframe. Based on these data, we used a meta-analysis statistic implemented by the R function “metarate” to estimate the expected fraction of undiagnosed patients that become newly diagnosable per year through growth of knowledge about Mendelian diseases. This rate was estimated as 6.74% (Supplementary Methods and Supplementary Table S2).

To calculate the number of false positive notifications per diagnostic notification and total clinician burden, we assume the existence of a typical undiagnosed patients’ database containing *n* patients. We estimate the average number of false positive notifications per patient per month *f* as the number of false positive notifications (FPs) per patient per month during the reanalysis experiment, calculated as 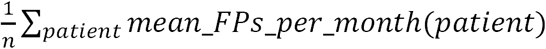 *mean_FPs_per_month(patient)*. Further, we estimate the fraction *p* of diagnosable patients who receive a diagnostic notification by automatic reanalysis as the fraction of diagnosable test patients who receive a diagnostic notification in the reanalysis experiment timeframe. Based on these estimates, the expected annual number of diagnostic notifications equals 6.74% · *n · p* and the expected annual number of false positive notifications equals 12 · *f* · *n*. Thus, given a scenario in which 6.74% of patients in an undiagnosed patients database become diagnosable within a year, the expected number of false positive notifications per diagnostic notification equals 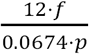 and the total evaluation burden on clinicians, per patient per year, is 6.74% · *p + 12* · *f*.

#### Comparison of AMELIE-based reanalysis to a simple abstract-based approach

To estimate the efficiency gain of AMELIE-based reanalysis over a manual abstract-based reanalysis approach, we defined the 20 most cited Mendelian disease journals as the most-cited journals in the Human Gene Mutation Database (HGMD), which aims to comprehensively curate Mendelian disease-causing mutations from the primary literature^28^ (Supplementary Table S3 and Supplementary Methods). For each patient, we assembled a surveillance list of all articles mentioning at least one patient candidate causative gene in the 20 most cited Mendelian disease journals that were published between the start of the reanalysis experiment and the publication of the first diagnostic article for the patient. The first diagnostic article was contained in this surveillance list for 82-83% of patients (91 of 110 of singleton patients and 51 of 61 of trio patients). Consequently, we estimated the *efficiency gain of automatic reanalysis compared to tracking the 20 most cited Mendelian disease journals* for a patient equals the number of articles about any of the patient’s candidate causative genes in the 20 most cited journals about Mendelian disease until publication of the first diagnostic article divided by the number of AMELIE-based automatic reanalysis notifications for the patient.

#### Notification threshold calibration

The automatic (global) reanalysis notification threshold can be adjusted to achieve high sensitivity (aiming for a large fraction of diagnosed patients), or high precision (aiming for a low number of false positives per diagnostic notification). We report the measures defined above for 3 differently calibrated notification thresholds: (a) a “*high-sensitivity*” notification threshold, in which the clinician receives diagnostic notifications for at least 80% of diagnosable patients, comparable in recall to tracking the top 20 journals above, at the lowest possible clinician burden, (b) a “*high-precision*” approach, in which at most 3 false positives per diagnostic notification are sent on average at the highest possible true positive rate, and (c) a “*minimal interruptions*” (even higher precision) approach, in which the majority of notifications sent are diagnostic, at the highest possible true positive rate.

## Results

Table 2 summarizes the outcomes of the reanalysis experiment. The fraction of diagnosed patients and total number of notifications per patient per year is shown in Figure 2. The automatic reanalysis timeline of three examples of singleton patients is presented in Table 3. Automatic reanalysis on singleton data could be calibrated for high sensitivity or high precision; achieving high sensitivity and precision simultaneously was possible with trio data. Both modes of operation resulted in between 86 and 893 times fewer abstracts to consider compared to manual reanalysis by tracking abstracts in the 20 most cited Mendelian disease journals.

**Table 2.**
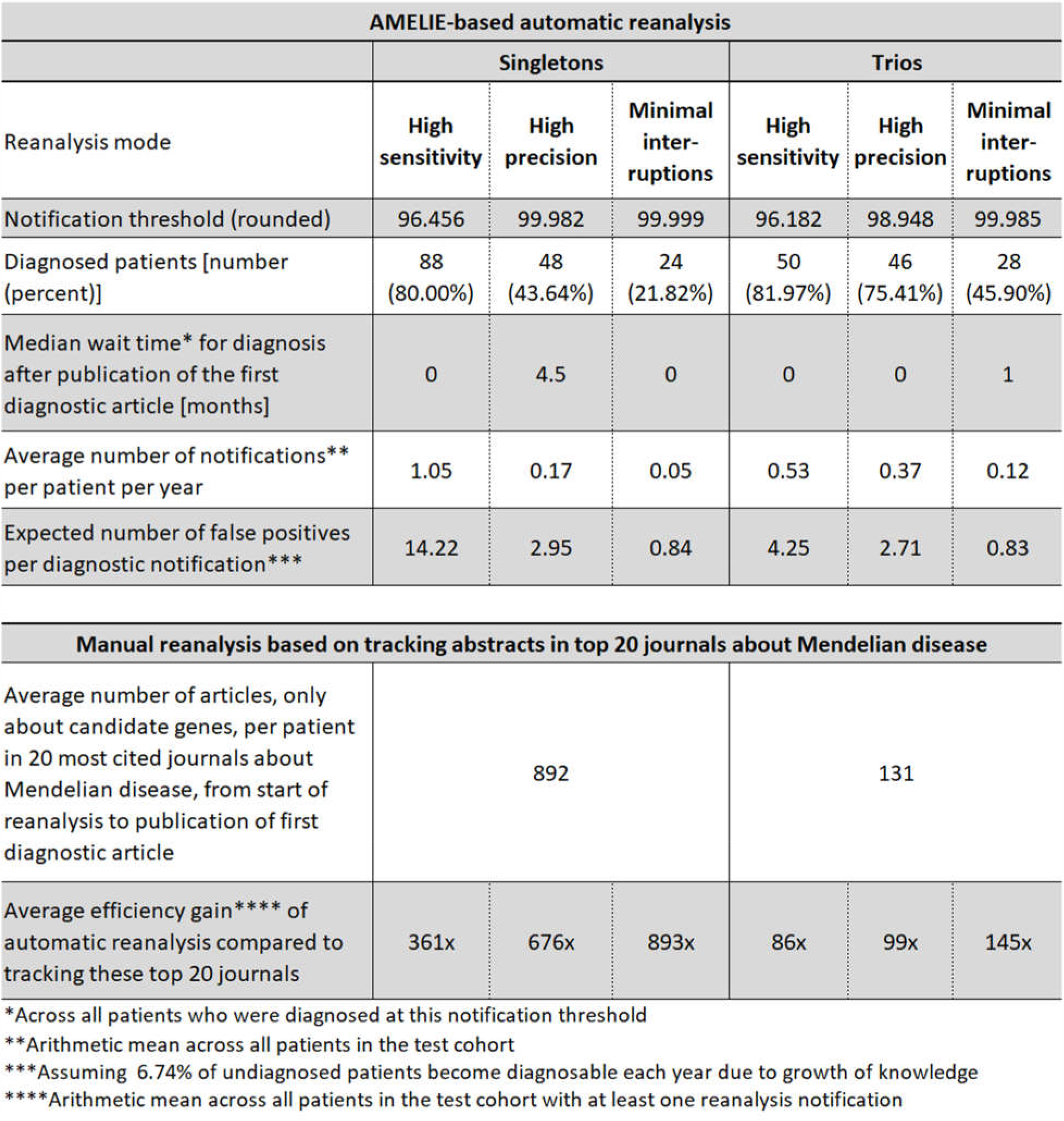
Reanalysis experiment outcomes.

**Table 3.** Automatic reanalysis notifications of three singleton patients, starting January 2012.

| Notification date*,** | Notification class | Reported Gene | Article title | Article first author and journal |
| --- | --- | --- | --- | --- |
| **Patient082. Diagnosis: "Metabolic encephalomyopathic crises, recurrent, with rhabdomyolysis, cardiac arrhythmias, and neurodegeneration".** |  |  |  |  |
| **Causative gene: TANGO2. First diagnosable\*: January 2016.** |  |  |  |  |
| (no notifications for 4 years, until: ) |  |  |  |  |
| January 2016 | Diagnostic | TANGO2 | Recurrent Muscle Weakness with Rhabdomyolysis, Metabolic Crises, and Cardiac Arrhythmia Due to Bi-allelic TANGO2 Mutations. | Lalani SR et al., American Journal of Human Genetics |
| **Patient032. Diagnosis: "Epileptic encephalopathy, childhood-onset".** |  |  |  |  |
| **Causative gene: CHD2. First diagnosable\*: May 2013.** |  |  |  |  |
| May 2013 | Missed | CHD2 | Targeted resequencing in epileptic encephalopathies identifies de novo mutations in CHD2 and SYNGAP1. | Carvill GL et al., Nature Genetics |
| November 2013 | Diagnostic | CHD2 | De novo loss-of-function mutations in CHD2 cause a fever-sensitive myoclonic epileptic encephalopathy sharing features with Dravet syndrome. | Suls A et al., American Journal of Human Genetics |
| **Patient073. Diagnosis: "Mental retardation, X-linked 102".** |  |  |  |  |
| **Causative gene: DDX3X. First diagnosable\*: August 2015.** |  |  |  |  |
| October 2012 | False positive | COL4A1 | Novel COL4A1 mutations cause cerebral small vessel disease by haploinsufficiency. | Lemmens R et al., Human Molecular Genetics |
| September 2014 | False positive | AFG3L2 | Deletion of AFG3L2 associated with spinocerebellar ataxia type 28 in the context of multiple genomic anomalies. | Myers KA et al., American Journal of Medical Genetics. Part A |
| August 2015 | Diagnostic | DDX3X | Mutations in DDX3X Are a Common Cause of Unexplained Intellectual Disability with Gender-Specific Effects on Wnt Signaling | Snijders Blok L et al., American Journal of Human Genetics |
| *PubMed publication date |  |  |  |  |
| **For singleton analysis at notification threshold 99.982 ("high precision" threshold) |  |  |  |  |

**Figure 2.**
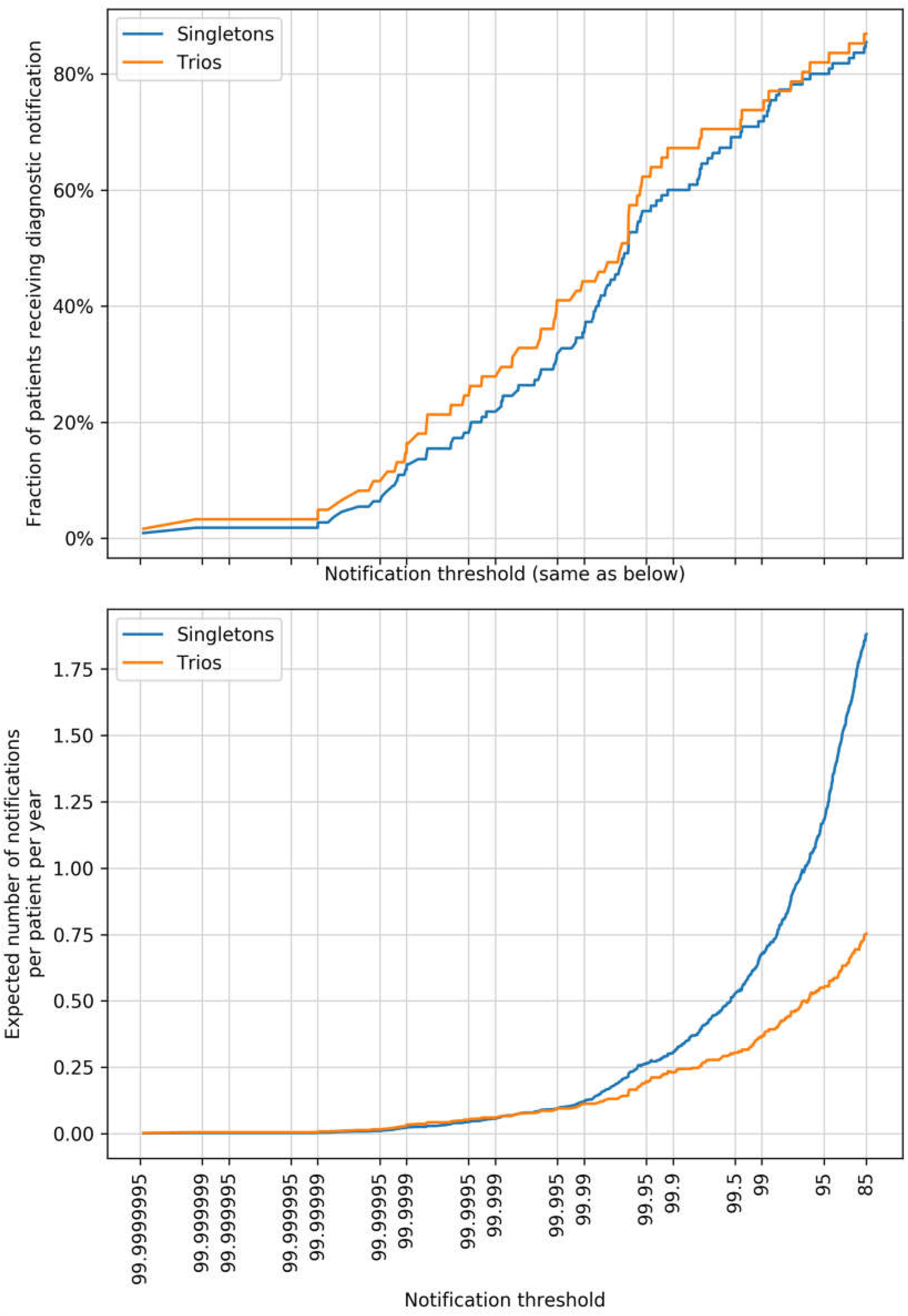
Fraction of diagnosed patients and average clinician burden per patient per year across notification thresholds. Both panels have the same x-axis so that matching values can read simultaneously from both. **(Upper panel)** The fraction of diagnosable test cohort patients who received a diagnostic notification (i.e., true positive) during the 6.5 year reanalysis experiment timeframe across notification thresholds. **(Lower panel)** The expected total number of notifications (or clinician burden) per patient per year across notification thresholds, including both diagnostic notifications and false positive notifications. For example, the system detects 80% of diagnosable singletons (trios) at the low burden of 1 (0.5) notification per patient per year.

### Singletons

We ran singleton analysis on all 110 patients. By manually tracking articles (only) about patient candidate causative genes in the 20 most cited Mendelian disease journals, clinicians would need to evaluate an average of 892 articles per diagnosable patient from the start of the reanalysis experiment until the publication of the first diagnostic article.

In contrast, our automatic reanalysis system is powerful enough to attain “high sensitivity”, where 80% of all diagnosable patients trigger a diagnostic notification, 58% of them immediately upon publication of the first diagnostic article, at an average of only 1.05 notification per patient per year (Figure 2 and Table 2). In “high precision” mode false positive notifications are reduced by 80%, while 44% of diagnosable singleton patients receive a diagnostic alert, at an average rate of only 0.17 notifications per patient per year. And in “minimal interruptions” mode, only 22% of diagnosable singleton patients receive a diagnostic notification, but the majority of notifications sent by the system are diagnostic, at a minimal 0.05 notifications per patient per year.

Thus, automatic reanalysis with the above notification thresholds for high sensitivity, high precision, or minimal interruptions, requires following up on 361-893 times fewer articles compared to manual reanalysis surveillance overall, amounting to only a couple of article alerts per patient.

### Trios

In the case of manual reanalysis for our 61 trio patients, clinicians would examine an average of 131 articles about candidate causative genes per patient by tracking abstracts in the 20 most cited Mendelian disease journals from start of the reanalysis experiment to the publication of the first diagnostic article.

In contrast, automatic trio reanalysis in “high sensitivity” mode resulted in an 82% diagnosis rate, at 0.53 notifications per patient per year, or half the clinician burden of comparable singleton reanalysis. “High precision” mode was very similar, resulting in over 75% of diagnoses. And in “minimal interruptions” mode, the diagnosis rate was still 46% of diagnosable patients with the majority of notifications leading to diagnosis, at an impressive 0.12 notifications per patient per year.

Thus, automatic reanalysis as presented here requires following up on 86-145 times fewer articles per patient compared to manual reanalysis by tracking abstracts in the 20 most cited Mendelian disease journals.

### Web portal

We have launched a web portal containing a working implementation of AMELIE analysis^21^ followed by automatic reanalysis at https://amelie.stanford.edu. The updated website is trained on current PubMed (as opposed to 2011 in above experiment), and it performs daily literature updates by automatically parsing and classifying newly indexed PubMed entries, downloading full text of relevant articles, and inserting extracted knowledge from full-text articles into the AMELIE knowledgebase. For demonstration purposes users can sign up for individual accounts and enable automatic reanalysis notifications (delivered by email) for selected patients at user-defined notification thresholds. Customizable singleton and trio variant filtering based on gnomAD variant frequency data^29^ is supported.

## Discussion

We present here a retrospective analysis of an automatic reanalysis framework on both singleton patients and trios diagnosed with Mendelian disorders over the span of over six years. We showed that automatic reanalysis can already be used to reveal diagnoses for patients with suspected Mendelian disease who could not be previously diagnosed at a very acceptable notification burden, while requiring dramatically less work of clinicians as compared to manual reanalysis. By simply tracking abstracts pertaining to patient candidate causative genes in the 20 most cited Mendelian disease journals, clinicians have to review hundreds of articles per diagnosable patient from the start of our reanalysis experiment to diagnosis.

In 2016 we were among the first to publish on the value of reanalysis^6^. From 40 cases we were able to diagnose 4. This 10% yield (on cases accumulated over multiple years) has since held up for a great number of similar studies by other groups over their undiagnosed patients. Here our sample size is bigger, and we expect it to be similarly representative of continuous patient reanalysis at under 1 notification per patient per year. Moreover, AMELIE’s “time machine” performance here was obtained while training *only* on 2011 data, not long after next generation sequencing became available in the clinic. It should be seen as a lower bound on AMELIE’s actual performance, as the AMELIE web portal is trained on nearly a decade of additional years of accumulated knowledge. Performance would further improve should the conservative expected rate of reanalysis diagnoses per year we estimate at 6.7% be higher.

A mass of sequenced but undiagnosed patients is already accruing^17^. CLIA-certified exome data production now costs only a few hundred dollars. A wave of data – millions of sequenced patients^3^, and tens of thousands of articles on Mendelian disease genes^21^ – is coming the way of fewer than a thousand clinical laboratory geneticists in the U.S.^30^ and their peers worldwide. Germline exome and genome sequencing data, in contrast to results from many other diagnostic tests, do not expire. As our knowledge about disease-causing genetic variation constantly grows, manual reinterpretation of patient sequencing data can at best be done periodically. In Mendelian diagnosis alone, a substantial 70% of cases will not be diagnosed at initial analysis^5^, and yet, as estimated here, a meaningful ∼6.7% will become diagnosable with each subsequent year that passes on new knowledge alone. This accumulating load will greatly weigh on any interpretation service. Automation, as we show, can realize the promise of continuous reanalysis and timely diagnosis for all, and will be essential to handle the incoming flood of healthcare data and insights.

Our AMELIE-based reanalysis framework has limitations, catching only 80% of diagnosable cases even in high sensitivity mode. But what diagnoses it finds, it offers with an efficiency gain of ∼100-1000-fold over the – unsustainable – current standard of manual curation. Importantly, our system does not replace clinicians, but rather augments their capabilities. If a medical institute or lab devotes a certain number of work hours to re/analysis, a small fraction of this time should be devoted to resolving our system’s notifications. The remainder can certainly be spent on more open-ended explorations, and all lessons learned (both inside and outside the system) can be incorporated to make such resident clinical support systems better and better over time.

Traditionally, patient cases are most often reassessed at the time of a new clinical encounter. The rapid accumulation of medical knowledge pressures this paradigm as the significance of one’s health record can change dramatically between visits. On any given day, a patient may become diagnosable and a portion of such diagnoses are expected to be immediately actionable. At the same time, logistical and cost constraints currently prevent the regular reanalysis of many patient cases following non-diagnostic sequencing. Together with automated phenotype extraction tools from the electronic medical record, like ClinPhen^22^, AMELIE demonstrates the potential of a scalable means of regular reanalysis for undiagnosed patients, which can also encompass emerging incidentals. This has implications for the care of patients with undiagnosed genetic disease and more broadly. The promise of efficient, continuous, automated identification of latent, actionable diagnoses in patient data has the potential to significantly improve health outcomes across care settings.

## Data Availability

A portion of the data we use is available from EGA. Another portion is of consented Stanford or UDN patients. Some of the latter can be shared while respecting consent conditions.

## Acknowledgments

We would like to thank Erich Weiler for continuous support and guidance. We thank the members of the Bejerano lab for technical advice and helpful discussions. We thank Victoria Wang, Max Haeussler, Mark E. Diekhans, Natalie T. Deuitch, and Laura E. Hayward for helpful input. We thank Elijah Kravets, Julia Buckingham and Kirstie MacMillan for study coordination. We thank the European Genome-Phenome Archive^27^ (EGA) and the Deciphering Developmental Disorders (DDD) project^25^ for data sharing. The DDD study presents independent research commissioned by the Health Innovation Challenge Fund [grant HICF-1009-003], a parallel funding partnership between the Wellcome Trust and the Department of Health, and the Wellcome Trust Sanger Institute [grant WT098051]. The views expressed in this publication are those of the author(s) and not necessarily those of the Wellcome Trust or the Department of Health. The study has UK Research Ethics Committee approval (10/H0305/83, granted by the Cambridge South REC, and GEN/284/12 granted by the Republic of Ireland REC). Deidentified DDD data was obtained through EGA. The research team acknowledges the support of the National Institute for Health Research, through the Comprehensive Clinical Research Network. The authors would like to thank the Genome Aggregation Database (gnomAD) and the groups that provided exome and genome variant data to this resource. A full list of contributing groups can be found at http://gnomad.broadinstitute.org/about. UDN data were obtained directly from the UDN.

## Author contributions

JB and GB designed the study and analyzed the results. JB and ES implemented the text mining software, website and associated databases. EEB verified diagnostic articles for the purposes of the reanalysis experiment. CAD and KAJ processed patient data. JNK, DB, SM, JAMA, SN, CGP, JDC, RH, JMS, JBK, JAR, PM, DRA, VS, EAW, CME, EAA, MTW, and UDN provided curated patient data. PDS and DNC curated HGMD. JAB provided guidance on clinical aspects of study design, testing set construction and interpretation of results. JB, JAB, and GB wrote the manuscript. All authors commented on and approved the manuscript. GB guided the study.

